# Patterns of COVID-19 pandemic dynamics following deployment of a broad national immunization program

**DOI:** 10.1101/2021.02.08.21251325

**Authors:** Hagai Rossman, Smadar Shilo, Tomer Meir, Malka Gorfine, Uri Shalit, Eran Segal

**Affiliations:** Department of Computer Science and Applied Mathematics, Weizmann Institute of Science, Rehovot, Israel; Department of Molecular Cell Biology, Weizmann Institute of Science, Rehovot, Israel; Pediatric Diabetes Clinic, Institute of Diabetes, Endocrinology and Metabolism, Rambam Health Care Campus, Haifa, Israel; Department of Statistics and Operations Research, Tel Aviv University, Ramat Aviv, Israel; Technion - Israel Institute of Technology, Haifa, Israel

## Abstract

Studies on the real-life impact of the BNT162b2 vaccine, recently authorized for the prevention of coronavirus disease 2019 (COVID-19), are urgently needed. Here, we analysed the temporal dynamics of the number of new COVID-19 cases and hospitalization in Israel following a rapid vaccination campaign initiated on December 20th, 2020. We conducted a retrospective descriptive analysis of data originating from the Israeli Ministry of Health (MOH) from March 2020 to February 2021. In order to distill the possible effect of the vaccinations from other factors, including a third lockdown imposed in Israel on January 2021, we compared the time-dependent changes in number of COVID-19 cases and hospitalizations between (1) individuals aged 60 years and older, eligible to receive the vaccine earlier, and younger age groups; (2) the latest lockdown (which was imposed in parallel to the vaccine rollout) versus the previous lockdown, imposed on September 2020; (3) early-vaccinated cities compared to late-vaccinated cities; and (4) early-vaccinated geographical statistical areas (GSAs) compared to late-vaccinated GSAs. In mid-January, the number of COVID-19 cases and hospitalization started to decline, with a larger and earlier decrease among older individuals, followed by younger age groups, by the order in which they were prioritized for vaccination. This fast and early decline in older individuals was more evident in early-vaccinated compared to late-vaccinated cities. Such a pattern was not observed in the previous lockdown. Our analysis demonstrates evidence for the real-life impact of a national vaccination campaign in Israel on the pandemic dynamics. We believe that our findings have major public health implications in the struggle against the COVID-19 pandemic, including the public ’s perception of the need for and benefit of nationwide vaccination campaigns. More studies aimed at assessing the effectiveness and impact of vaccination both on the individual and on the population level, with longer followup, are needed.

## Introduction

An effective and safe vaccination campaign is urgently needed to halt the rapid spread of *Severe acute respiratory syndrome coronavirus 2* (SARS-CoV-2) infections and the resulting disease, Coronavirus disease 2019 (Covid-19). The BNT162b2 vaccine, developed by BioNTech in cooperation with Pfizer, is a lipid nucleoside-modified RNA (modRNA) encoding the SARS-CoV-2 full-length spike ^1^. On December 11, 2020, the Food and Drug Administration (FDA) issued an Emergency Use Authorization (EUA) for emergency use of the vaccine for the prevention of COVID-19 ^2^. Results from a phase III randomized placebo-controlled trials demonstrated that a two-dose regimen in a 21 days interval conferred 95% protection against laboratory-confirmed Covid-19 infection in individuals 16 years of age or older ^3^.

On December 20th 2020, Israel launched its COVID-19 vaccination campaign ^4^, in which BNT162b2 vaccines were administered. In Israel, the public health system is comprised of only four health maintenance organizations (HMOs) and vaccinations were widely available for the country ’s entire population, according to the following prioritization, determined by the Israeli ministry of health (MOH): During the early phases of the distribution process, individuals considered as being at high risk for COVID-19 were given a priority in vaccination, including individuals older than 60 years old, nursing home residents, healthcare workers, and patients with severe comorbidities. The vaccination campaign was further expanded for individuals aged 55 years old ^5^ and 40 years old ^6^ or older, on January 12th and January 19th 2021 respectively. On the 21st of January, pupils aged 16-18 years old were also prioritized for vaccination. On the 28th of January, the vaccination campaign expanded to those aged 35 and older ^7^. On the 4th of February, all individuals aged 16 years old and older were eligible to receive the vaccine. However, the HMOs were still instructed to focus their efforts on those aged 50 years old and older ^8^. Individuals with a history of severe allergic reactions to the vaccine components, individuals who recovered from COVID-19 or those younger than 16 years of age (with the exception of children with severe chronic diseases) are not eligible to receive the vaccine as of February 24th.

The national vaccination campaign has led Israel to be the country with the highest rate of vaccinated individuals per capita, with 48.8%, 34% and 7.5% of the population having received the first or the second vaccine dose, or recovered from COVID-19, respectively as of February 24th, 2021, or 68.7%, 48% and 8% respectively, taking into account population older than 16 years old - the population currently eligible for vaccination. In parallel, during the early weeks of the vaccination campaign, the number of cases and hospitalized patients has rapidly increased, along with the local emergence of the B117 variant ^9^, leading the government to impose a third lockdown on 8th of January 2021. This lockdown was gradually eased starting on the 7th of February 2021.

When evaluating the effects of vaccines there are two complementary forms of evaluation: one is measuring the direct effects of the vaccine on the vaccinated individual (termed “vaccine effectiveness”, VE), and the other is measuring the overall impact of the vaccination programme on an entire population (termed “vaccine impact”, VI) ^10^. On the individual level, preliminary studies from two of the largest Israeli HMO ’s have attempted to estimate the real-life effectiveness of the vaccine. The first reported an efficacy of 51% for the first dose after 13-24 days ^11^ while the second reported an efficacy of 46% and 92% following 14-20 days from the first dose and 7 or more days from the second doses of the vaccine respectively ^12^. Another report estimated that the vaccine effectiveness is above 95% 3-4 weeks after the second dose ^13^. However, to our knowledge, very few studies ^14^ thus far have analysed the impact of the vaccination campaign on the patterns of pandemic dynamics at the population level. As Israel is one of the first countries to implement a vaccination campaign on this magnitude, we believe that this quantification may be of major interest for many countries worldwide.

## Methods

A retrospective analysis on data was conducted on data originating from the Israeli MOH from March 2020 to February 2021. The data included information on age, sex, date of positive SARS-CoV-2 polymerase chain reaction (PCR) test, date of hospitalization, clinical state during hospitalization and date of death or recovery for each individual. Data on national vaccination is available online (http://data.gov.il/dataset/covid-19/) and includes the number of daily vaccine doses, separated to first and second doses, administered in each city by age groups (a scale of 10 years). Recovered individuals were included in the vaccinated group in all of the analysis, as previous studies analyzing the antibody responses following recovery showed persistent antibody titers against the SARS-CoV-2 spike protein for several months after infection ^15^. Notably, throughout the study period, these individuals were not eligible to receive the vaccine. A total of 3,210,200 individuals vaccinated with both doses of the vaccine and 711,949 recovered individuals were included in the analysis.

The temporal changes in weekly numbers of several clinical measures were analysed including positive COVID-19 cases, percentage of positive tests, hospitalized patients, and hospitalized patients in a severe state. Covid-19 cases were identified by a positive SARS-CoV-2 polymerase chain reaction (PCR) test. Classification of the hospitalization severity was based on the following clinical criteria, applied on 13 of July 2020 by the Israeli MOH^16^ based on NIH ^17^ and WHO^18^ definitions: *Mild illness -* individuals who have any of the various signs and symptoms of COVID 19 (e.g., fever, cough, malaise, loss of taste and smell); *Moderate illness* - individuals who have evidence of pneumonia by a clinical assessment or imaging; *Severe illness -* individuals who have respiratory rate >30 breaths per minute, SpO2 <93% on room air at sea level, or ratio of arterial partial pressure of oxygen to fraction of inspired oxygen (PaO2/FiO2) <300 mmHg and *Ventilated/Critical -* individuals with respiratory failure who require ventilation (invasive or non-invasive), multiorgan dysfunction or shock. In this work, we denote all patients in a severe case or worse as severe (including ventilated and critical patients).

In order to distill the impact of the vaccination campaign from other factors that may influence COVID-19 morbidity and mortality, including a third lockdown imposed in Israel during this time period, the following comparisons of the dynamic of the clinical measures mentioned above were performed: First, we compared between individuals aged 60 years and older, who were the population prioritized to receive the vaccine earlier, and younger individuals, who were prioritized according to the MOH guidelines (Fig. 1). Second, we compared between the decline in the number of cases and hospitalizations observed following the initiation of the second national lockdown imposed by the Israeli government on 18th of September 2020, with the decline observed following the initiation of the third national lockdown, imposed while the vaccination program was in place, on the 8th of January 2021 (Fig 3.).

**Fig. 1.**
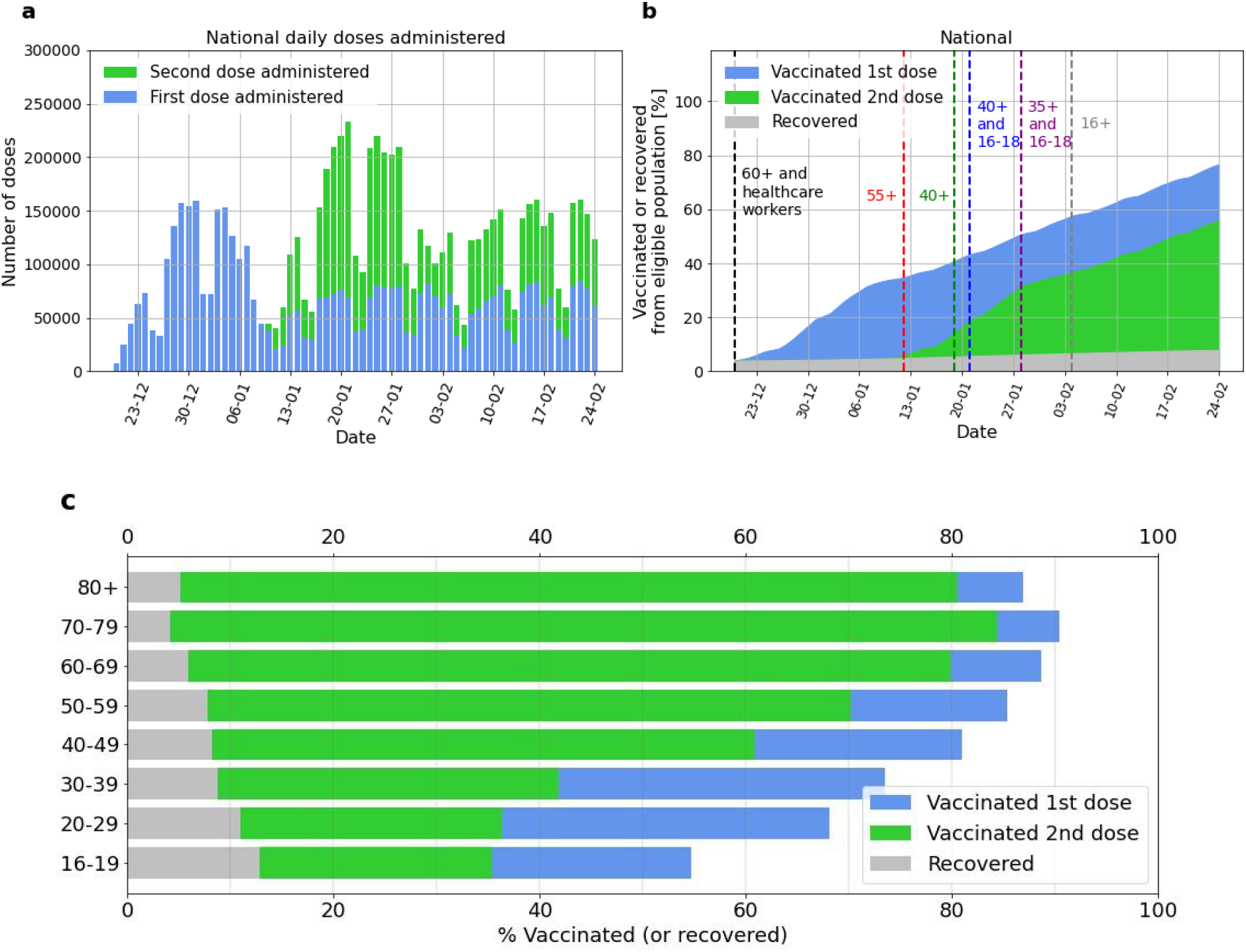
**a**. Daily national number of administered vaccination doses, first dose (blue bars) and second dose (green bars). **b**. Cumulative percentage of the national population recovered or vaccinated. Vaccinated population that received the first dose is shown in blue; the vaccinated population that received the second dose is shown in green. Recovered population is shown in gray. Times at which age groups were formally eligible for vaccination are shown as dashed vertical lines. **c**. Vaccinated or recovered percentage per age group on Feb 24th 2021.

Third, cities with a high percentage of individuals who were vaccinated early were compared to cities with a low percentage: For every city with more than 5,000 residents aged 60 and older the percentage of these individuals who received the first dose of the vaccine was calculated. *Early-vaccinated cities* were defined as the top 10 cities with the highest percentage of individuals older than 60 years that were either vaccinated or recovered from COVID-19 by the 10th of January 2021 (10 cities with a total population of 641,276). *Late-vaccinated cities* were defined as the 10 bottom cities with the lowest percentage of individuals older than 60 years vaccinated by the 10th of January 2021 (10 cities with a total population of 1,845,759), see Fig. S5.

Finally, data with a higher geographical-resolution termed geographical statistical areas (GSAs) were analysed. The GSAs are small, relatively homogeneous intra-city units defined by the Israeli Bureau of Statistics, with an average of 3,000 residents, within cities with more than 10,000 inhabitants. Data on vaccination for the GSAs was available without age group separation, though the national vaccination policy described earlier still holds information regarding the timing of each age-group ’s vaccination. Out of 1385 GSAs, 1148 had more than 500 residents aged 60 years and older. Out of these 1148 GSAs, the 400 with the highest vaccination rate by January 10th (3 weeks after the beginning of the vaccine drive), 2021 were denoted as *early-vaccinated GSAs* and the 400 with the lowest vaccination rate by January 10th, 2021 were denoted as *late-vaccinated GSAs*.

## Results

Starting on the 20th of December 2020, the initiation of the vaccination campaign, the number of vaccines administered per day began at approximately 50,000, quickly rose to over 150,000 by December 24th 2020, and reached a maximum of 231,010 on January 21st 2021 (Fig 1). Vaccinations per day for each age group are shown in the supplementary material (SM) (Fig S1). By January 7th, almost 70% of the population over 60 years old have already been vaccinated (1st dose) or recovered, increasing gradually to 88.5% (1st dose) and 81% (both doses) by February 24th 2021.

The temporal changes in the number of new COVID-19 cases and hospitalizations in Israel from December 18th to February 24th are summarised in Fig 2 and 4. Several days after the initiation of the lockdown on January 8th and the beginning of the administration of the second vaccine doses on January 10th, the number of new COVID-19 cases aged 60 years old and older reached a peak. This peak was followed by a peak in mild, moderate or severe hospitalization of individuals, and a later peak in severe hospitalization of the same age group. Between January 15 and February 24 the number of new cases and hospitalizations in this age group declined. As several potential factors other than the vaccines may have influenced this decline, the following analyses were done, aimed at estimating the potential role of the vaccination campaign.

**Fig. 2.**
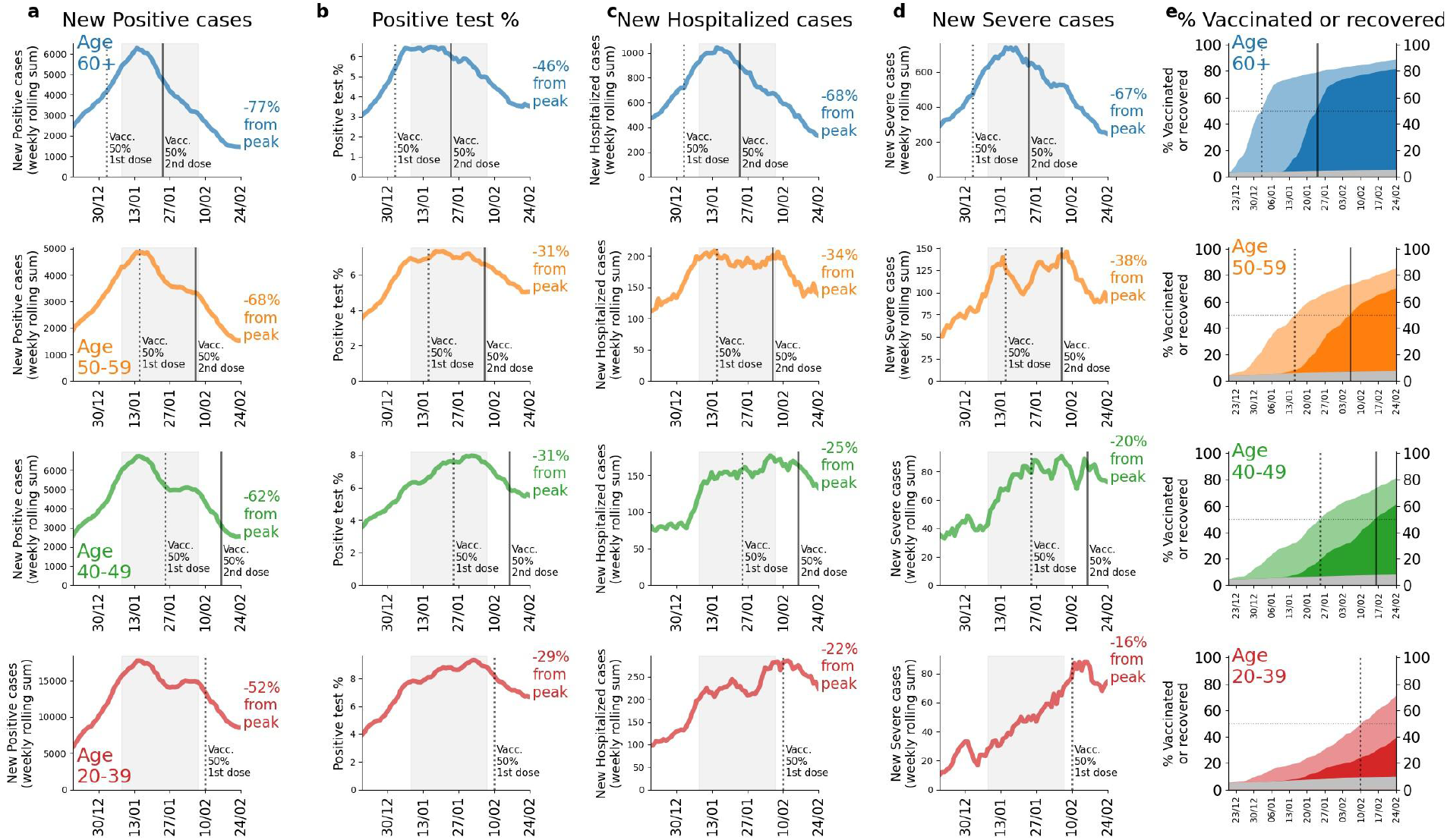
Dynamics of national clinical measures for different age groups: First row (blue) - aged over 60 years old, second row (orange) - aged 50-59, third row (green) - aged 40-49, fourth row (red) - aged 20-39. Columns: **a**. Rolling weekly sum of new positive PCR cases **b**. Percent of PCR tests which come out positive **c**. Rolling weekly sum of new hospitalizations **d**. Rolling weekly sum of new severe hospitalizations **e**. Cumulative vaccinated population percentage per age group. Light color represents the population percentage vaccinated with a first dose ; Dark coloring represents the population percentage per age group that has been vaccinated with a second dose; Grey colouring represents the recovered population percentage per age group (not eligible for vaccination). In all figures, the date at which 50% of the age group population has been vaccinated by first dose or recovered is shown as a black dotted vertical line, and the date at which 50% of the population has been vaccinated with a second dose or recovered is shown as a black vertical line. In columns A-D, the lockdown time period is displayed as a gray filling and the percentage reduction with respect to the peak value of the last date (February 24th) is presented at the tail of each curve.

**Fig. 3.**
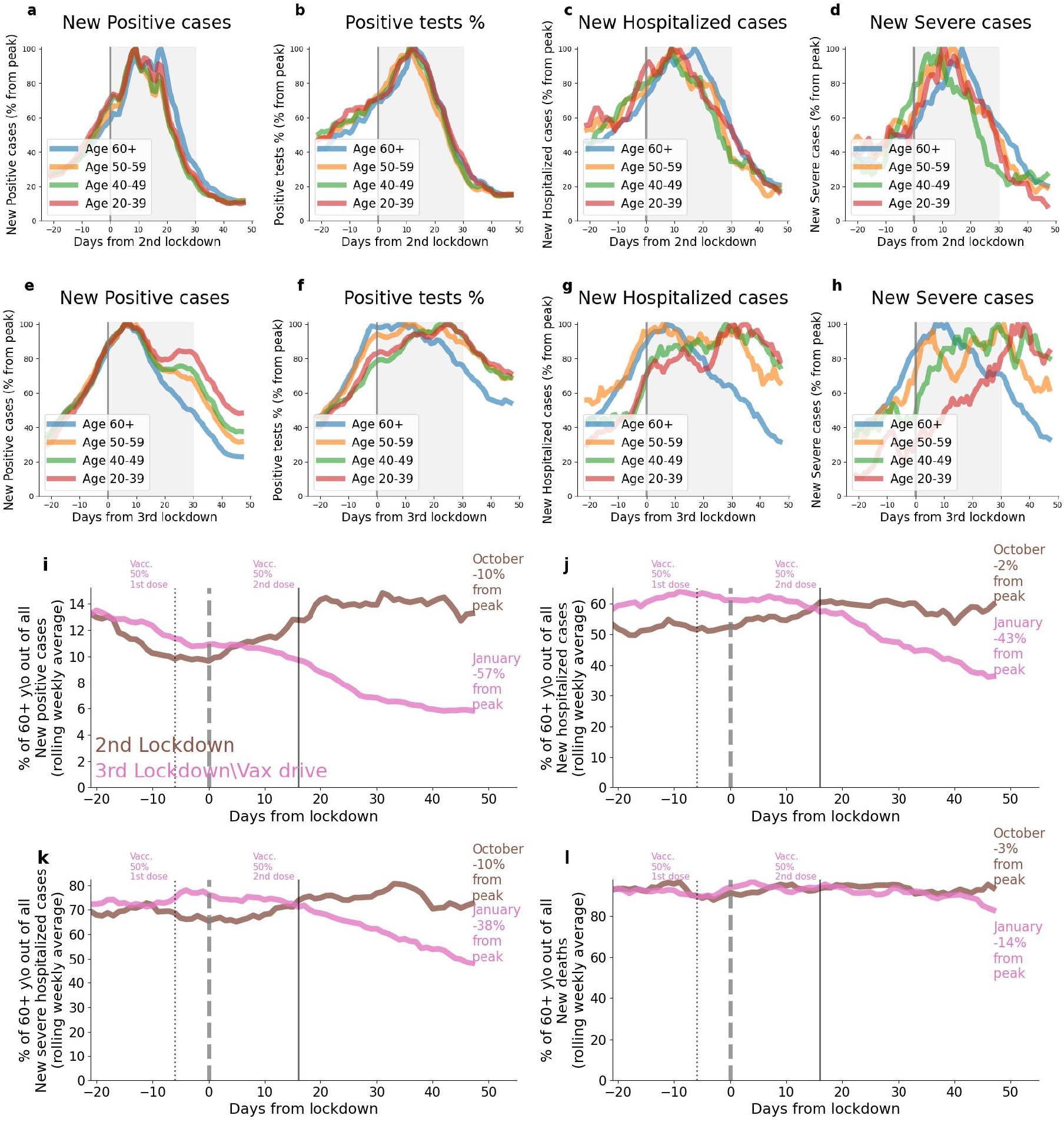
Comparison between age groups 20-39 (red), 40-49 (green), 50-59 (orange) and over 60 (blue) years old in **a**. Percent drop of new positive cases from peak value at the time period around the second lockdown **b**. Percent drop of the percent of PCR tests which are positive, relative to the peak at the time period around the second lockdown **c**. Percent drop of new hospitalizations from the peak at the time period around the second lockdown **d**. Percent drop of new severe hospitalizations from the peak at the time period around the second lockdown **e**. Percent drop of new positive cases from peak value at the time period around the third lockdown **f**. Percent drop of the percent of PCR tests which are positive, relative to the peak at the time period around the third lockdown **g**. Percent drop of new hospitalizations from the peak at the time period around the third lockdown **h**. Percent drop of new severe hospitalizations from the peak at the time period around the third lockdown. In **a-d** “Day 0” represents the second lockdown start date, September 18th 2020. In **e-h** “Day 0” represents the third lockdown start date, January 8th 2021. In **a-h** the lockdowns time period is displayed as a grey filled area. **i-l** show the percent of age group 60+ years old out of the population of **i**. New positive cases **j**. New hospitalizations **k**. New severe hospitalizations **l**. New deaths. The percentage around the second lockdown is shown in brown, and the percentage around the third lockdown is shown in pink. The day in which 50% of the population has received the first dose or recovered is displayed as a dotted black vertical line, and the day in which 50% of the population has received the second dose or recovered is displayed as a black vertical line (relevant with respect to the third lockdown only).

**Fig. 4.**
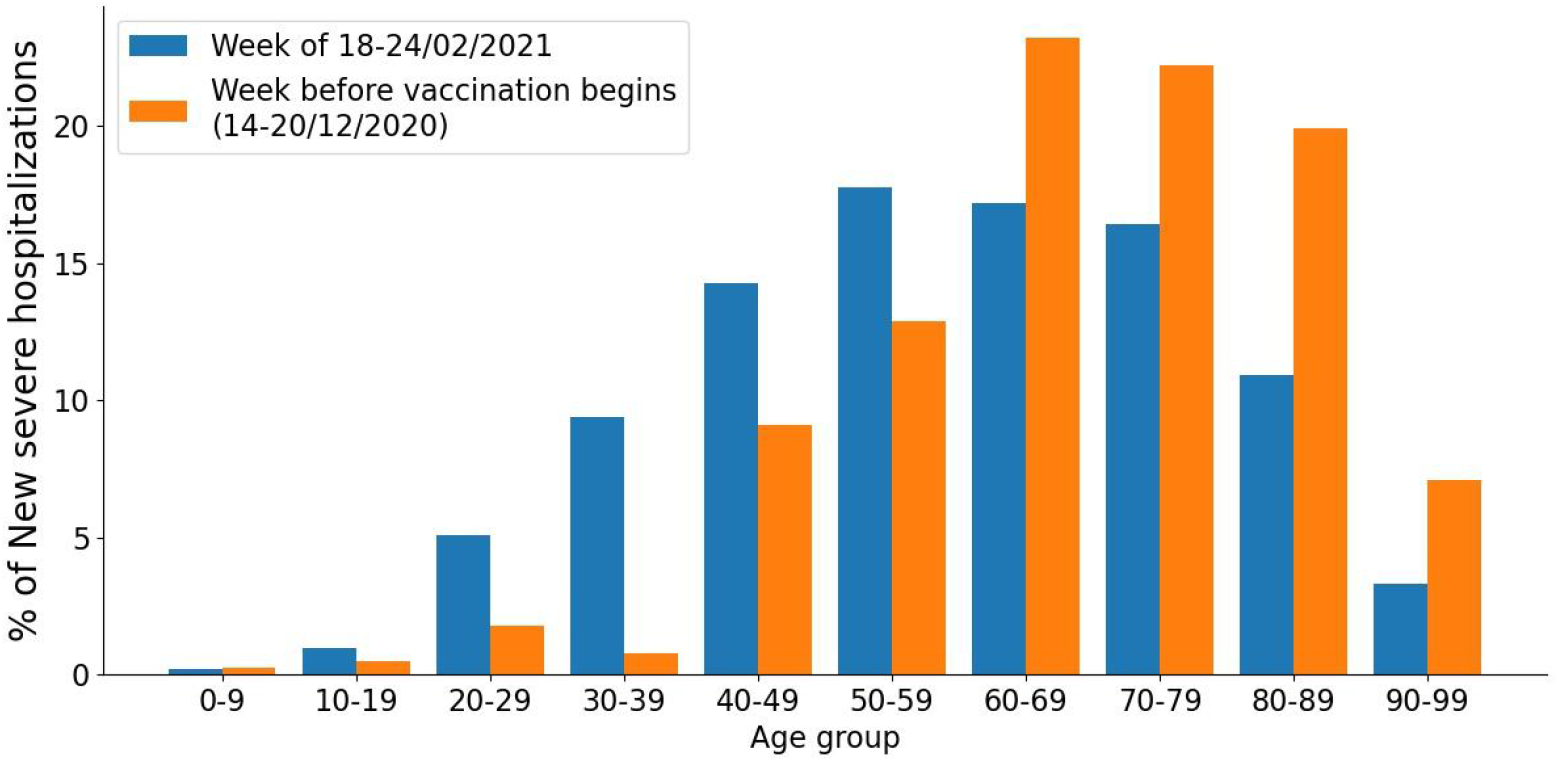
Percentage of each age group of all new severe hospitalizations at two different weeks: The week before the vaccination campaign initiated (orange) and the week of February 18th-24th (blue).

**Fig 5.**
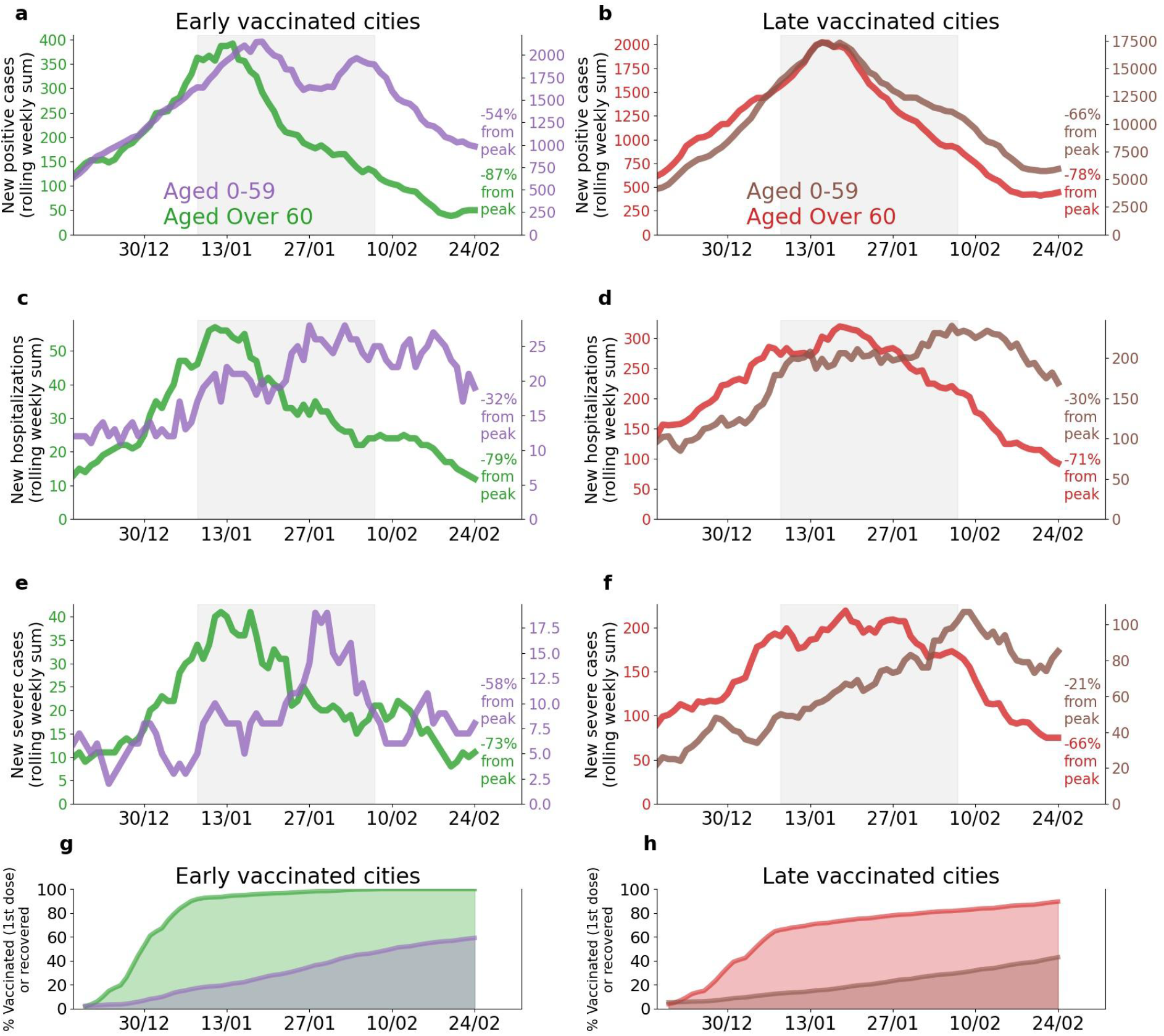
Comparison between age groups 0-59 years old and 60+ years old from cities with most of the population vaccinated early and cities with most of the population vaccinated late. All figures are for the time period of December 18th 2020 to February 24th 2021. In figures a-f the lockdown time period is shown as gray filling. Note: Figures a-f are presented with 2 different y-axis scales in order to highlight relative within-population trends. Age group 0-59 is shown as a purple line in a,c,e and as a brown line in b,d,f. Age group 60+ is shown as a green line in a,c,e and as a red line in b,d,f. **a**. Rolling weekly sum of new positive cases in early-vaccinated cities. **b**. Rolling weekly sum of new positive cases in late-vaccinated cities. **c**. Rolling weekly sum of new mild, moderate or severe hospitalizations in early-vaccinated cities. **d**. Rolling weekly sum of new mild, moderate or severe hospitalizations in late-vaccinated cities. **e**. Rolling weekly sum of new severe hospitalizations in early-vaccinated cities. **f**. Rolling weekly sum of new severe hospitalizations in late-vaccinated cities. **g**. Cumulative percentage of the population recovered or vaccinated (1st dose) in early-vaccinated cities. Age group 60+ is shown as a green line, age group 0-59 is shown as a purple line. **h**. Cumulative percentage of the population recovered or vaccinated (1st dose) in late-vaccinated cities. Age group 60+ is shown as a red line and age group 0-59 is shown as a brown line.

We first compared individuals 60 years, who were prioritized to receive the vaccine earlier, with younger individuals (Fig. 2). Notably, the decrease in number of cases and hospitalizations was larger and earlier in older individuals compared to younger individuals. Moreover, the decrease in these clinical measures was according to the order of real-life vaccination rate (Fig S3), which was guided by the prioritization made by the Israeli MOH. For example, a decrease of 46% versus 29% in the percentage of positive tests and 68% versus 22% in hospitalizations compared to the peak was observed in individuals 60 years and older compared to individuals aged 20-39 years. This is also evident in a significant shift towards younger ages in the distribution of ages of severe COVID-19 patients (Fig.4). In order to allow youth to attend their school exams, adolescents (age 16-18 years) were also prioritized relatively early in the vaccination campaign. Many young adults (age 18-21 years old) serving in the Israel Defense Forces were also prioritized. Notably, COVID-19 cases and the positivity rates of diagnostic tests dropped more rapidly in this group (age 16-21 years old) compared to similar age groups which were not vaccinated yet (13-15 and 22-23 years old), (Fig S4).

Fig 3 presents a set of comparisons of clinical measures between two time periods: a period following the second national lockdown imposed by the Israeli government on 18th of September 2020 (Fig. 3 a-d), and recent times following the third national lockdown, imposed on the 8th of January 2021 (Fig. 3 a-d). Notably, while all clinical measures had almost the same dynamics for all age groups during the first period (2nd lockdown), these dynamics differed significantly across age groups in the second period (3rd lockdown+vaccine drive), where a larger and earlier decline in older individuals (age above 60 years old) compared to younger individuals was apparent.

The analysis between *early-vaccinated* cities and *late-vaccinated* cities (see Methods) revealed a larger and earlier decrease in the number of COVID-19 cases and hospitalizations of individuals 60 years old and older in cities vaccinated early compared to late-vaccinated cities. For example, in early-vaccinated cities, there was a decrease of 89% in cases and of 79% in severe hospitalizations compared to peak values, while in late-vaccinated cities a smaller decrease of 78% in cases and 66% in severe hospitalizations was observed. Finally, in order to obtain a higher geographic resolution we compared between *early-vaccinated* GSAs and *late-vaccinated* GSAs (see Methods) (Fig. 4). This analysis revealed similar findings as the city level analysis.

**Table 1.** Vaccination status and decrease in cases on Feb. 24th with respect to the peak values. A summary table of Fig 2.

|  | a | b | c | d | e |  |  |
| --- | --- | --- | --- | --- | --- | --- | --- |
| Age group, years | New Positive cases | Positive Tests Percentage | New Hospitalized Cases | New Severe Cases | Vaccinated - First Dose [%] | Vaccinated - Second Dose [%] | Recovered [%] |
| 60+ | -77% | -46% | -68% | -67% | 83.7% | 76.4% | 4.7% |
| 50-59 | -68% | -31% | -34% | -38% | 77.6% | 62.4% | 7.4% |
| 40-49 | -62% | -31% | -25% | -20% | 72.8% | 52.6% | 7.8% |
| 20-39 | -52% | -29% | -22% | -16% | 60.8% | 29.0% | 9.4% |

**Table 2.** Percent change of COVID-19 cases and hospitalizations in early- and late-vaccinated cities, calculated with respect to the peak values. A summary table of Fig 5.

|  | Age group, years | change from the peak |  |
| --- | --- | --- | --- |
|  |  | Early cities | Late cities |
| New cases | 0-59 | -54% | -66% |
| New cases | 60+ | -89% | -78% |
| New Hospitalizations: |  |  |  |
| Mild, Moderate or Severe | 0-59 | -38% | -30% |
| Mild, Moderate or Severe | 60+ | -82% | -71% |
| Severe | 0-59 | -67% | -21% |
| Severe | 60+ | -79% | -66% |

## Discussion

Here we show early signs for the effect of a national vaccination campaign in Israel on the pandemic dynamics. Our analysis revealed that a little over two months after the initiation of the vaccination campaign, with 85% of individuals older than 60 years old already vaccinated with 2 doses (February 24th, 2021), there was an approximately 77% drop in cases, 46% in positive test percentage, 68% in hospitalizations and 67% in severe hospitalizations, compared to peak values. Although multiple other factors besides the vaccines may have influenced these results, several observations suggest that these patterns might be driven to a considerable degree by the vaccines. First, the decline in the measures above is greater in older individuals who were prioritized to receive the vaccine earlier, with consecutive drops observed in younger age groups later, according to the order of vaccine prioritization, including earlier drops in some young groups (16-21 years old) prioritised over older groups (21-35 years old). Second, the effect was greater in cities and GSAs where a higher fraction of individuals were vaccinated earlier. Finally, we did not observe a similar pattern of a larger and faster decline of cases and hospitalizations in older individuals during the previous lockdown imposed in Israel, in which all clinical measures had very similar dynamics in all age groups (Fig 3, SM. Figs S3, S4, S5, S6, S9 & S10).

Notably, although previous reports, both in a clinical trial setting ^3,11^ and on real-life data ^12^ have indicated that efficacy of the vaccine is already evident after the first dose, the improvement in the number of hospitalized patients in Israel has been dramatically evident approximately 3-4 weeks following the vaccination campaign. We believe that this has several reasons. First, the real-life effectiveness may be different from the efficacy reported in the clinical trial due to the logistics of refrigeration, storage, transportation and on-site administration of the vaccines in real-world settings and during a rapid deployment campaign may have been imperfect, thus lowering effectiveness. Second, the effect may be heterogeneous and population-dependent. For example, it is possible that older individuals, who were prioritized earlier in the vaccination campaign, may have a reduced or belated response to the vaccination due to a deterioration in both innate and adaptive immune function, also termed immunosenescence, as was previously shown for other vaccines ^19,20^. Third, it is possible that the effectiveness of the vaccine is reduced in light of the emergence of new and more violent viral strains, such as the B117 variant ^21^ (which is already prevalent in Israel at February 2021 according to the Israeli MOH), and the 501.V2 variant ^22^ which may be associated with an increased risk of death ^23^.

Even if the real-life effectiveness of the vaccine is comparable to the efficacy reported in the clinical trial, as shown by Dagan et al. ^12^, the impact of the vaccination programme on the population as a whole depends on additional factors, including the national vaccine coverage, vaccine allocation among different subgroups of individuals and social mixing of the different groups that may affect disease transmission through indirect effects. It is also possible that vaccinated individuals may alter their behaviour and decrease adherence to public health prevention guidance (e.g. physical distancing, face masks) thereby increasing viral transmission. Moreover, viral transmission may also occur in the vaccination sites themselves. The vaccination sites should be large and ventilated in order to decrease the probability of transmission on site. Finally, there is a clear trend where areas in Israel with higher infection rates and a lower socioeconomic status have lower vaccination rates, despite wide vaccine availability ^24^. This trend may also diminish or delay the overall effect of the campaign as those who are at a higher risk of being infected are less vaccinated. Further effort should therefore be made to encourage these populations to vaccinate and make the vaccines even more easily accessible for them. We note that exact individual-level efficacy numbers cannot be deduced from our analysis, and that due to all of the above issues, our results may be consistent with efficacies that are either lower or greater than those reported in the original clinical trial.

Our study has several limitations. First and foremost, our study is an observational study as opposed to a randomized clinical trial and therefore causal effects are difficult to infer. Second, the comparison between the second and third lockdown may be influenced by factors such as the total number of COVID-19 cases in the beginning of each lockdown, testing policy, hospitalization policy and public compliance to the restrictions that may have changed with time. Similarly, differences between cities might be influenced by behavioral and social differences beyond the vaccines. However, none of these factors were likely to cause the different patterns observed in the different age groups reported here. Finally, the effects of the vaccination campaign observed here may be influenced by factors specific to Israel and its healthcare system, in which all citizens are mandated to join one of the official non-profit health insurance organizations. Financial and regional disparities in other health-care systems may impact the distribution and availability of vaccinations, thereby influencing the real life efficacy of the vaccines.

Overall, we show an analysis of large-scale real-world data from Israel demonstrating first signs of real-life effectiveness of a national vaccination campaign, Although our findings are preliminary, they have major public health implications for the struggle against the COVID-19 pandemic. More studies aimed at assessing the effectiveness of the vaccination on reducing the transmission of SARS-CoV-are needed both on the individual and on the population level with larger longitudinal followup and in additional populations.

## Data Availability

Data availability statement
The data that support the findings of this study originates from the Israel ministry of health.
National age-group level vaccination data:
https://data.gov.il/dataset/covid-19/resource/57410611-936c-49a6-ac3c-838171055b1f
Aggregated town-level and age groups vaccination data is available at:
https://data.gov.il/dataset/covid-19/resource/12c9045c-1bf4-478a-a9e1-1e876cc2e182
Aggregated GSA -level infection, hospitalization and vaccination data is available at:
https://data.gov.il/dataset/covid-19/resource/d07c0771-01a8-43b2-96cc-c6154e7fa9bd
Some restrictions apply to the availability of parts of the data used in the analysis and so are not publicly available.

## Data availability statement

The data that support the findings of this study originates from the Israel ministry of health. National age-group level vaccination data:

https://data.gov.il/dataset/covid-19/resource/57410611-936c-49a6-ac3c-838171055b1f Aggregated town-level and age groups vaccination data is available at: https://data.gov.il/dataset/covid-19/resource/12c9045c-1bf4-478a-a9e1-1e876cc2e182 Aggregated GSA -level infection, hospitalization and vaccination data is available at: https://data.gov.il/dataset/covid-19/resource/d07c0771-01a8-43b2-96cc-c6154e7fa9bd

Some restrictions apply to the availability of parts of the data used in the analysis and so are not publicly available.

## Code availability statement

Any relevant source code will be made available at: https://github.com/hrossman/Patterns-of-covid-19-pandemic-dynamics-following-deployment-of-a-broad-national-immunization-program

## Ethics Declarations

An exemption from institutional review board approval was determined by the Israeli Ministry of Health as part of an active epidemiological investigation, based on use of anonymous data only and no medical intervention.

## Authors contribution

H.R conceived the project, designed and conducted the analyses, interpreted the results and wrote the manuscript; S.S & T. M designed and conducted the analyses, interpreted the results and wrote the manuscript; M.G, U.S, E.S designed the analyses, interpreted the results, wrote the manuscript, supervised and conceived the project.

## Acknowledgments

We thank Meir Bruhim, Michal Goldberg and Eldad Sitbon for their contributions to our efforts.

## Supplementary appendix

**Fig S1.**
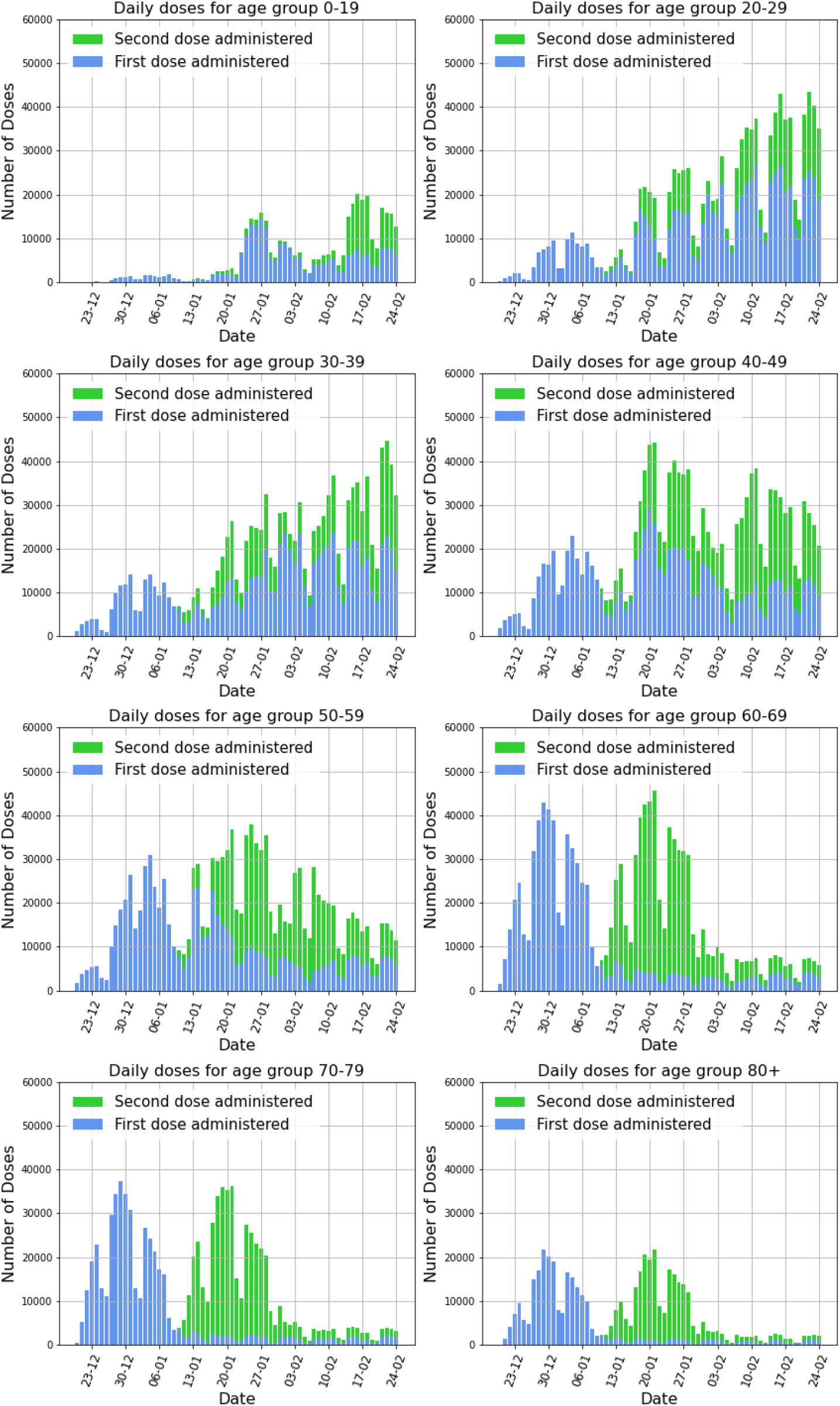
Daily number of administered vaccination doses by age groups, first dose (blue bars) and second dose (green bars).

**Fig S2.**
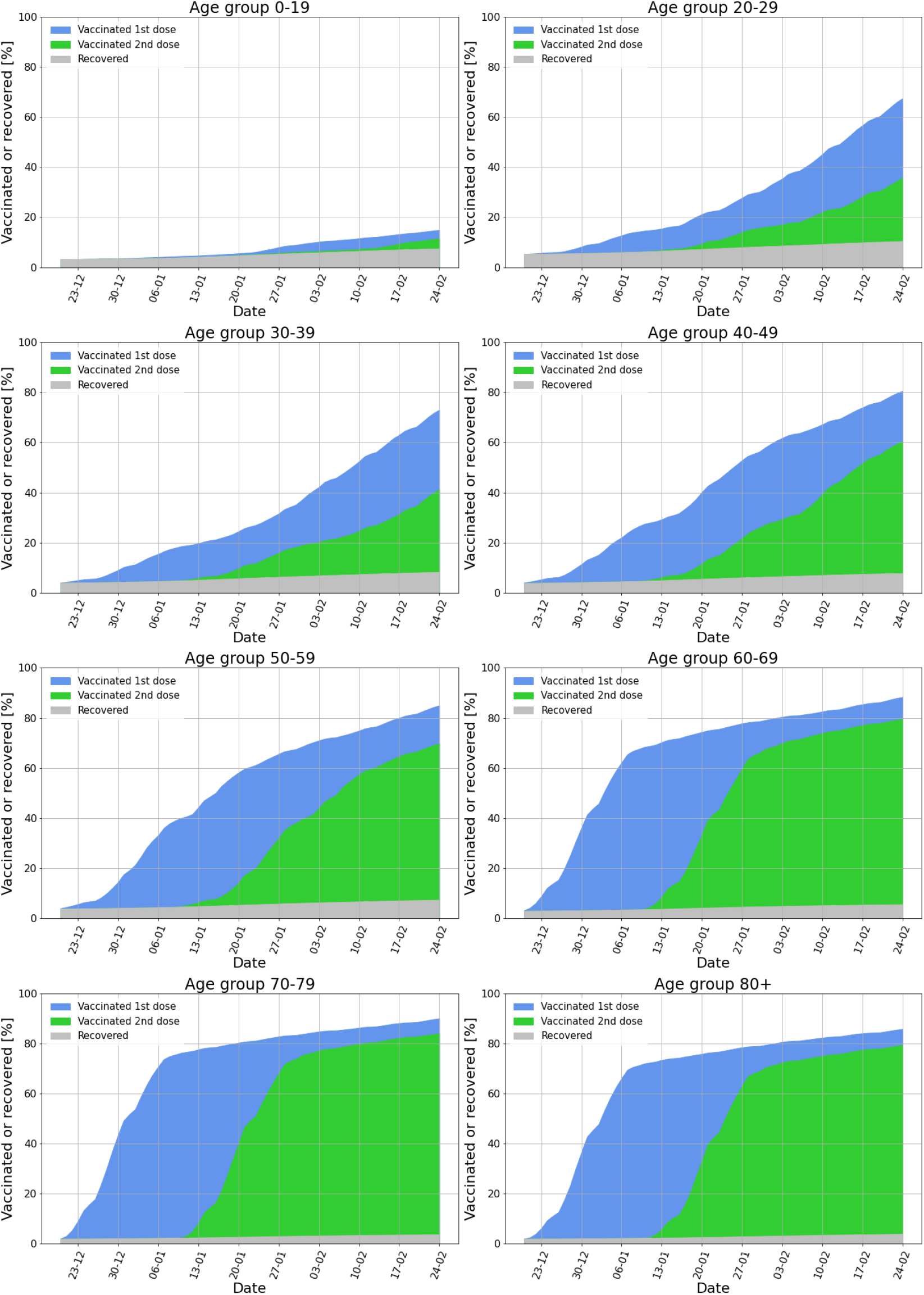
Cumulative percentage of each age-group recovered or vaccinated. Vaccinated population that received the first dose is shown in blue; and the vaccinated population that received the second dose is shown in green. Recovered population is shown in gray.

**Fig. S3.**
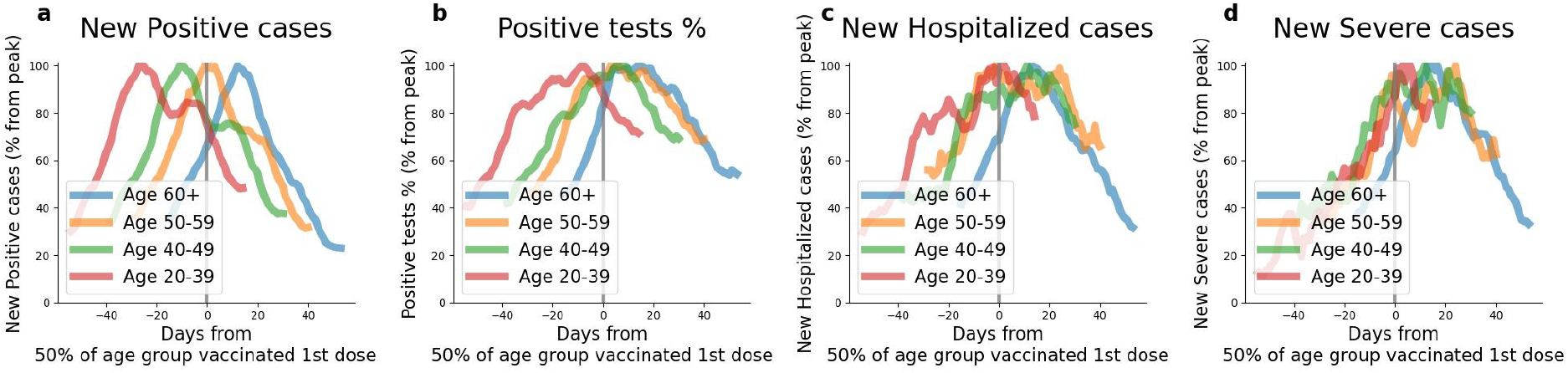
Comparison between age groups 20-39 (red), 40-49 (green), 50-59 (orange) and over 60 (blue) years old in **a**. Percent drop of new positive cases from peak value **b**. Percent from the peak of PCR tests positive results percentage **c**. Percent from the peak of new hospitalizations **d**. Percent from the peak of new severe cases. All curves are centered at the date in which 50% of the age group population has received the first dose or recovered.

**Fig. S4.**
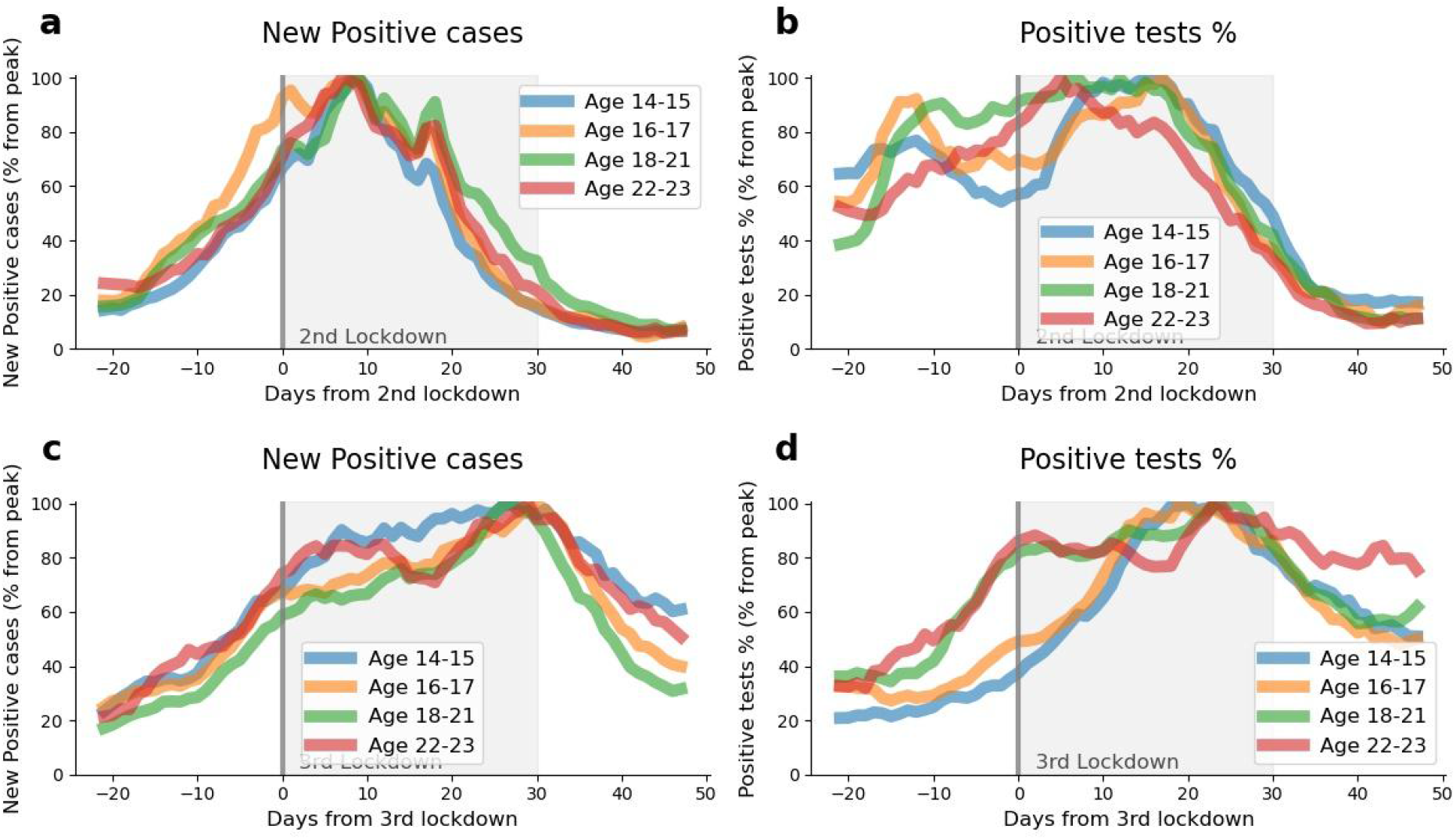
Comparison between age groups 14-15 (blue), 16-17 (orange), 18-21 (green) and 22-23 (red) years old in **a**. Percent drop of new positive cases from peak value at the time period around the second lockdown **b**. Percent from the peak of PCR tests positive results percentage at the time period around the second lockdown **c**. Percent drop of new positive cases from peak value at the time period around the third lockdown **d**. Percent from the peak of PCR tests positive results percentage at the time period around the third lockdown. In **a, b** “Day 0” represents the second lockdown start date, September 18th 2020. In **c, d** “Day 0” represents the third lockdown start date, January 8th 2021. In all figures the lockdown time period is displayed as gray filling.

**Fig S5.**
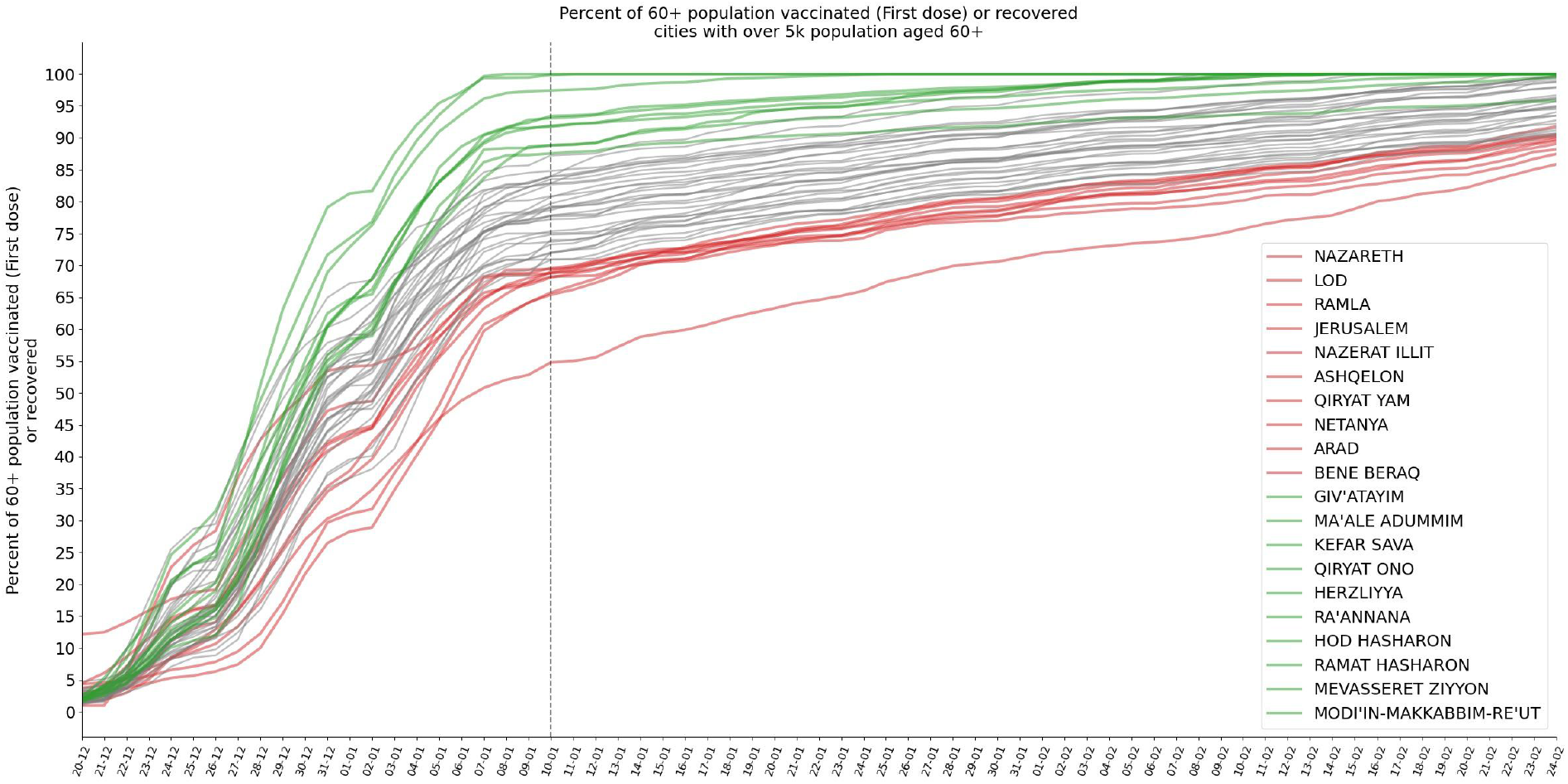
Cumulative percentage of vaccinated population from age group 60+, in cities with more than 5,000 residents in this age group. Early-vaccinated cities are shown as green lines, late-vaccinated cities are shown as red lines. Other cities are shown as gray lines.

**Fig S6.**
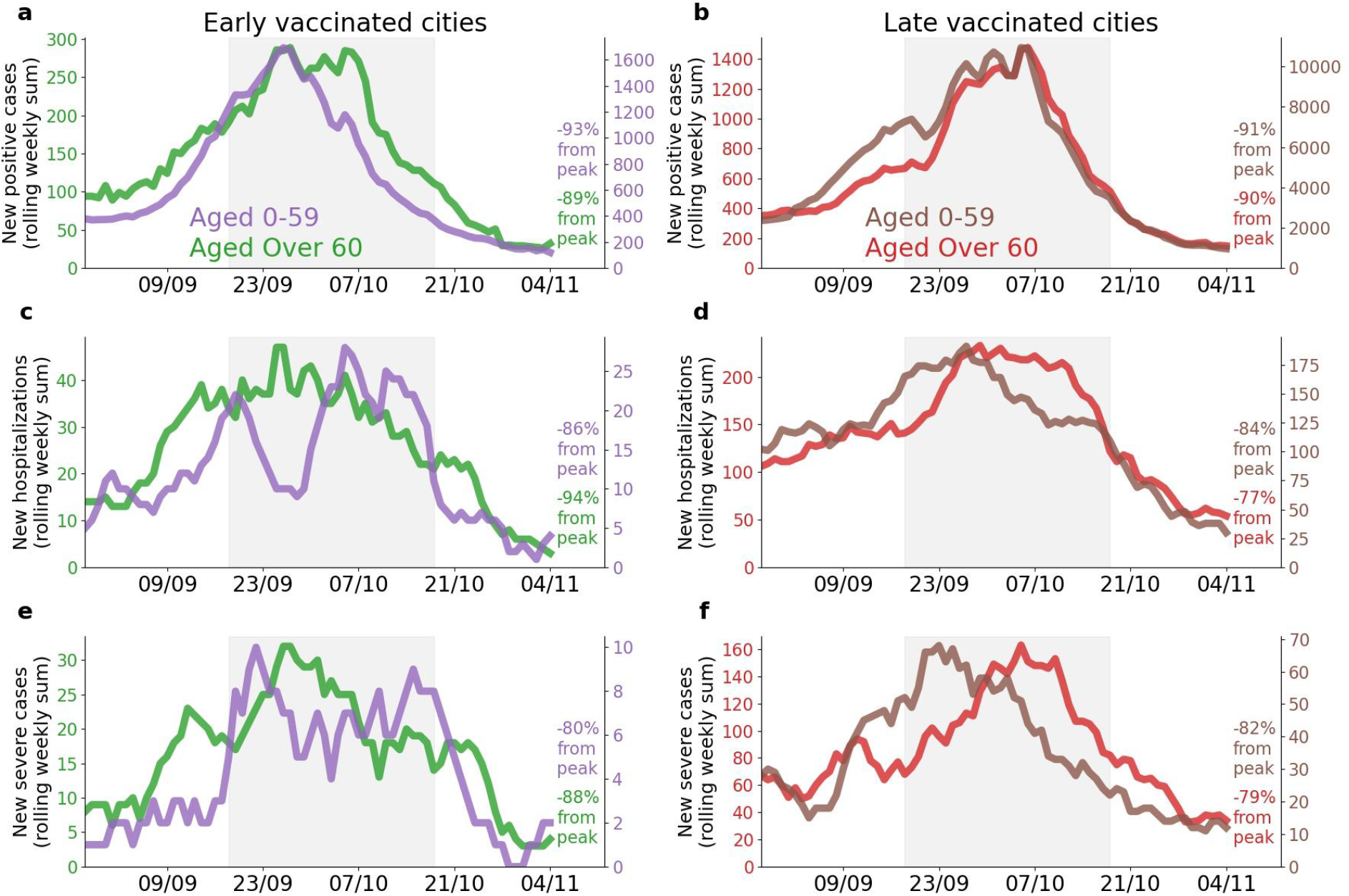
Comparison between age groups 0-59 years old and 60+ years old from cities with most of the population vaccinated early and cities with most of the population vaccinated late. In all figures the lockdown time period is shown as gray filling. Note: Figures a-f are presented with different y-axis scales in order to highlight relative within-population trends. Age group 0-59 is shown as a purple line in a,c,e and as a brown line in b,d,f. Age group 60+ is shown as a green line in a,c,e and as a red line in b,d,f. **a**. Rolling weekly sum of new positive cases in early-vaccinated cities. **b**. Rolling weekly sum of new positive cases in late-vaccinated cities. **c**. Rolling weekly sum of new mild, moderate or severe hospitalizations in early-vaccinated cities. **d**. Rolling weekly sum of new mild, moderate or severe hospitalizations in late-vaccinated cities. **e**. Rolling weekly sum of new severe hospitalizations in early-vaccinated cities. **f**. Rolling weekly sum of new severe hospitalizations in late-vaccinated cities.

**Fig S7.**
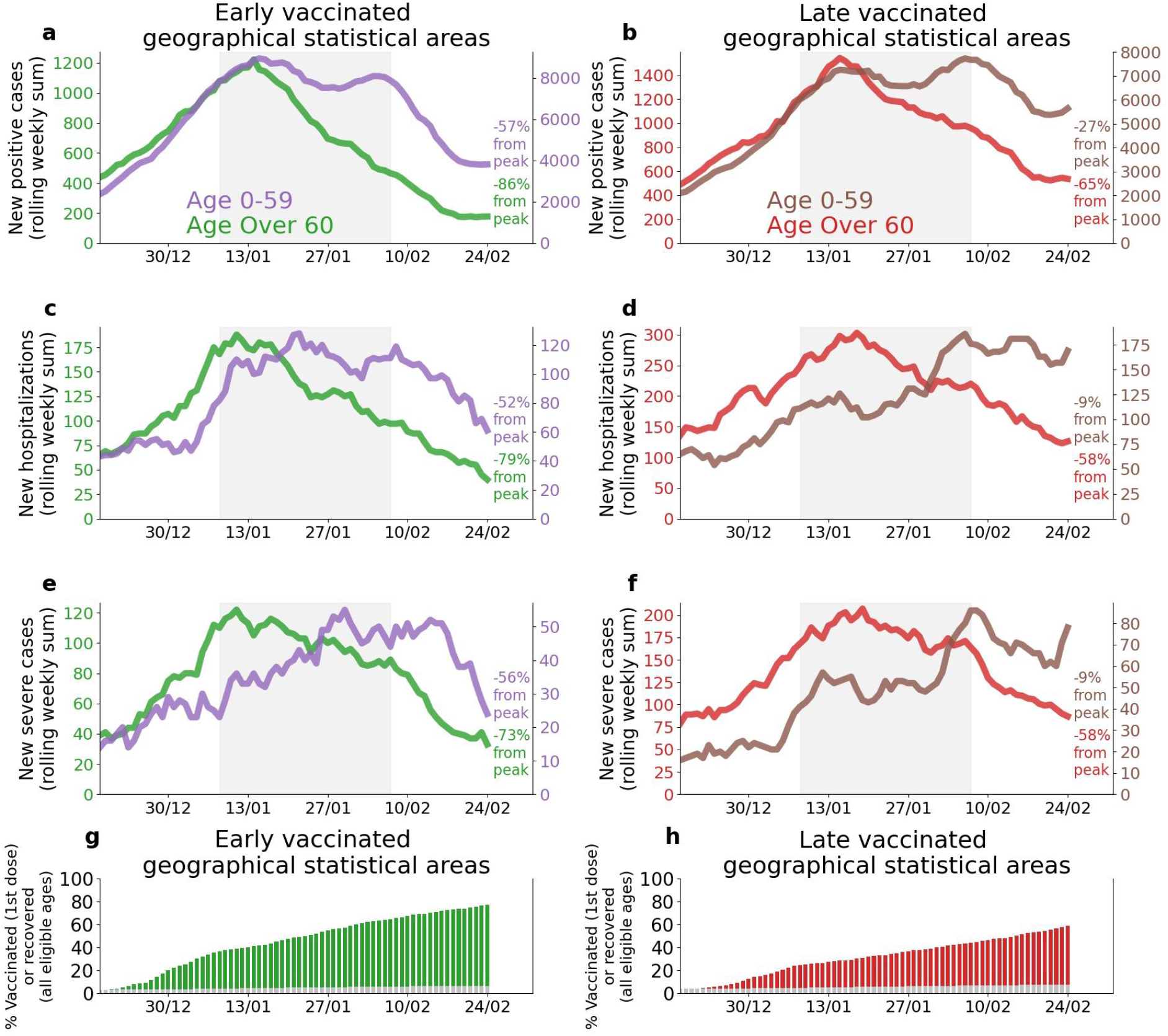
Comparison between age groups 0-59 years old and 60+ years old from early- and late-vaccinated GSAs. In all figures the third lockdown time period is shown as gray filling. Note: Figures a-f are presented with different y-axis scales in order to highlight relative within-population trends. Age group 0-59 is shown as a purple line in a,c,e and as a brown line in b,d,f. Age group 60+ is shown as a green line in a,c,e and as a red line in b,d,f. **a**. Rolling weekly sum of new positive cases in early-vaccinated GSAs. **b**. Rolling weekly sum of new positive cases in late-vaccinated GSAs. **c**. Rolling weekly sum of new mild, moderate or severe hospitalizations in early-vaccinated GSAs. **d**. Rolling weekly sum of new mild, moderate or severe hospitalizations in late-vaccinated GSAs. **e**. Rolling weekly sum of new severe hospitalizations in early-vaccinated GSAs. **f**. Rolling weekly sum of new severe hospitalizations in late-vaccinated GSAs. **g**. Percentage of individuals vaccinated or recovered in time in early-vaccinated GSAs. **h**. Percentage of individuals vaccinated or recovered in time in late-vaccinated GSAs.

**Table S1.**
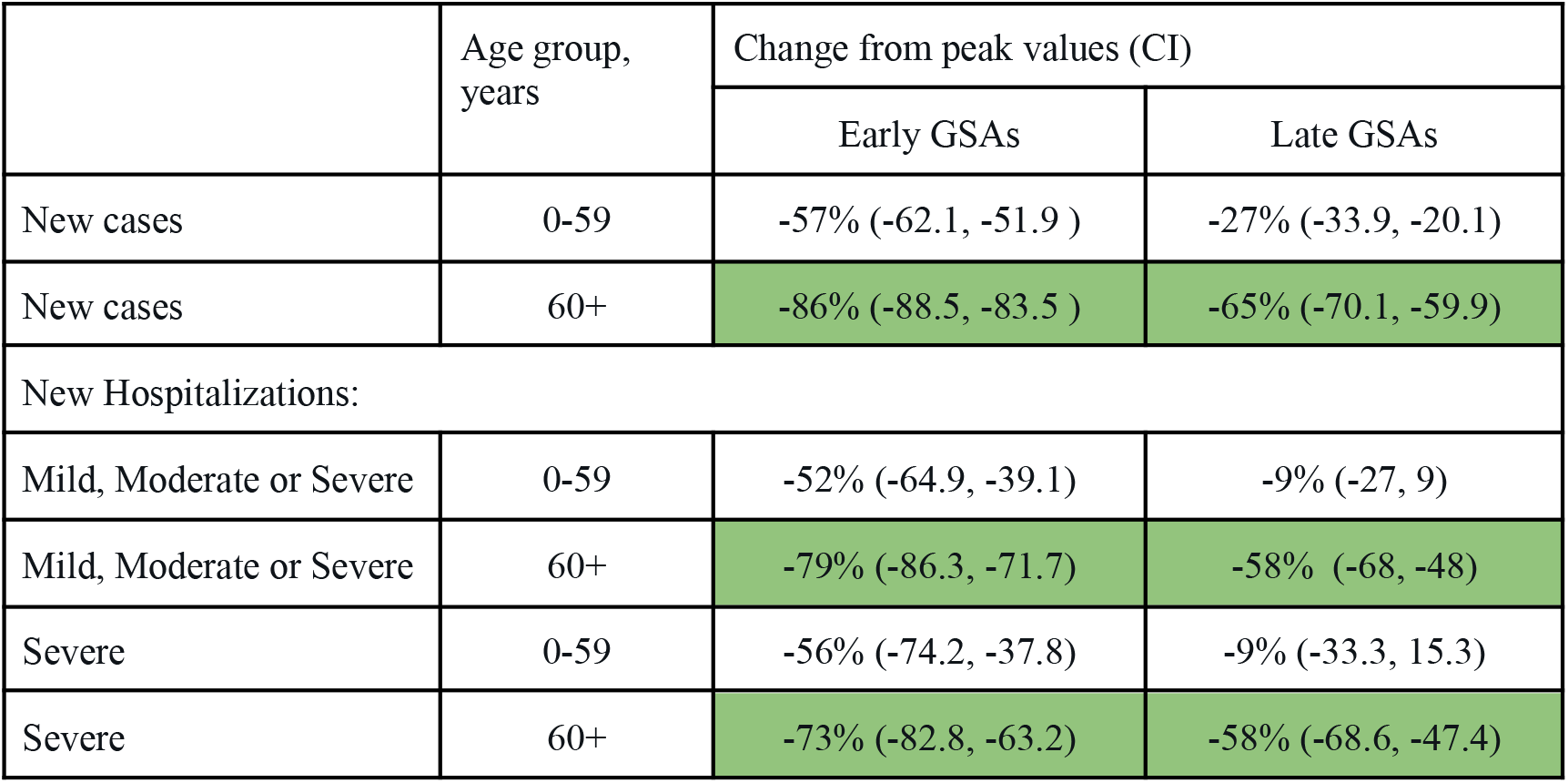
Percent change of COVID-19 cases and hospitalizations in early- and late-vaccinated Geographical Statistical Areas (GSAs), calculated with respect to the peak values. The GSA bootstrap standard errors were estimated by 500 bootstrap samples (each bootstrap sample consists of 400 late/early GSAs samples with replacement). Confidence intervals (CIs) were constructed based on the bootstrap standard errors.

|  | Age group,<br>years | Change from peak values (CI) |  |
| --- | --- | --- | --- |
|  |  | Early GSAs | Late GSAs |
| New cases | 0-59 | -57% (-62.1, -51.9 ) | -27% (-33.9, -20.1) |
| New cases | 60+ | -86% (-88.5, -83.5 ) | -65% (-70.1, -59.9) |
| New Hospitalizations: |  |  |  |
| Mild, Moderate or Severe | 0-59 | -52% (-64.9, -39.1) | -9% (-27, 9) |
| Mild, Moderate or Severe | 60+ | -79% (-86.3, -71.7) | -58% (-68, -48) |
| Severe | 0-59 | -56% (-74.2, -37.8) | -9% (-33.3, 15.3) |
| Severe | 60+ | -73% (-82.8, -63.2) | -58% (-68.6, -47.4) |

**Fig S8.**
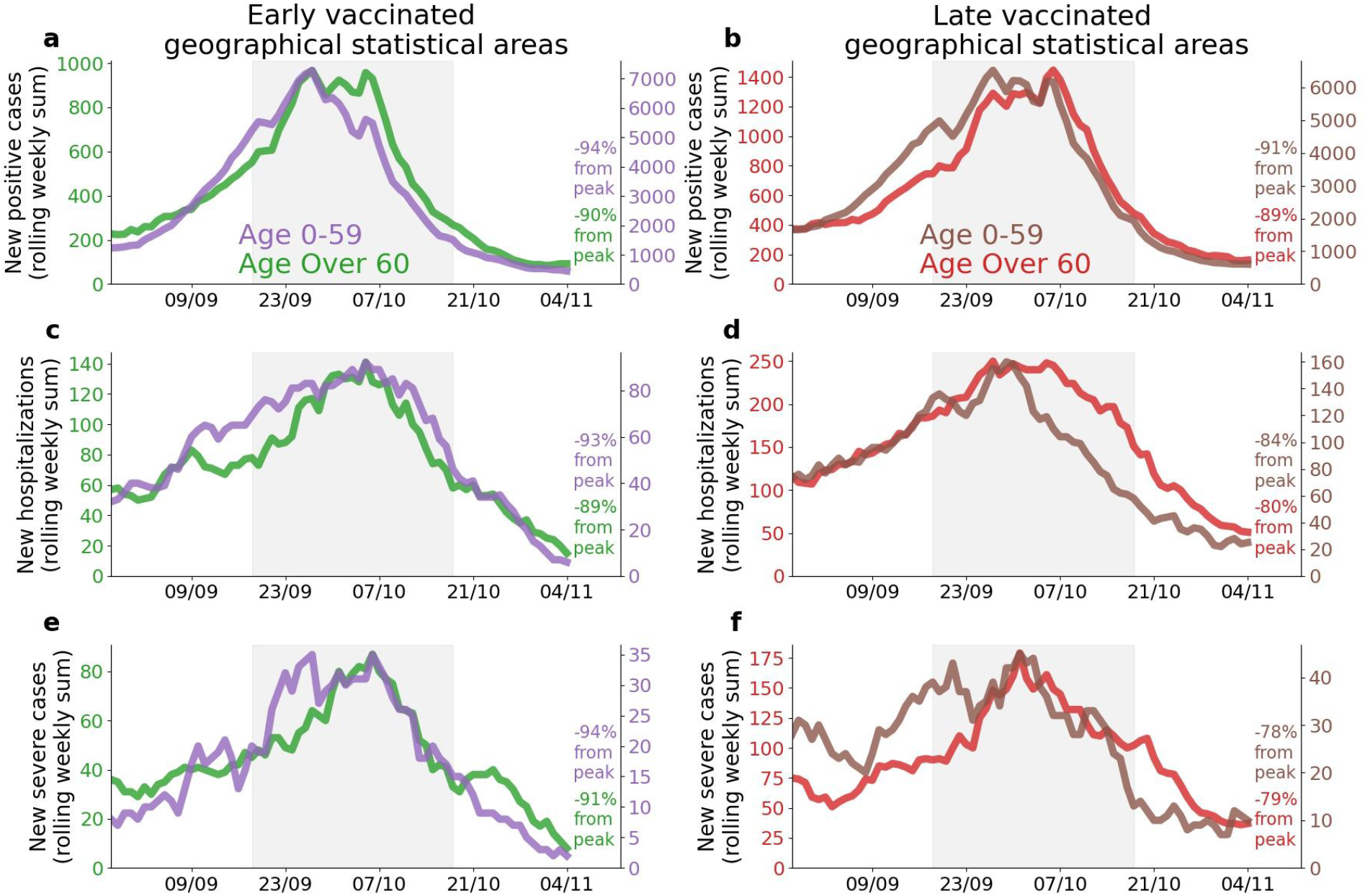
Comparison between age groups 0-59 years old and 60+ years old from geographical statistical areas (GSAs) with most of the population vaccinated early and GSAs with most of the population vaccinated late. In all figures the second lockdown is shown as gray filling. Note: Figures a-f are presented with different y-axis scales in order to highlight relative within-population trends. Age group 0-59 is shown as a purple line in a,c,e and as a brown line in b,d,f. Age group 60+ is shown as a green line in a,c,e and as a red line in b,d,f. **a**. Rolling weekly sum of new positive cases in early-vaccinated GSAs. **b**. Rolling weekly sum of new positive cases in late-vaccinated GSAs. **c**. Rolling weekly sum of new mild, moderate or severe hospitalizations in early-vaccinated GSAs. **d**. Rolling weekly sum of new mild, moderate or severe hospitalizations in late-vaccinated GSAs. **e**. Rolling weekly sum of new severe hospitalizations in early-vaccinated GSAs. **f**. Rolling weekly sum of new severe hospitalizations in late-vaccinated GSAs.

